# A Gaussian model for the time development of the Sars-Cov-2 corona pandemic disease. Predictions for Germany made on March 30, 2020

**DOI:** 10.1101/2020.03.31.20048942

**Authors:** R. Schlickeiser, F. Schlickeiser

## Abstract

For Germany it is predicted that the first wave of the corona pandemic disease reaches its maximum of new infections on April 11th, 2020 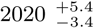 days with 90 percent confidence. With a delay of about 7 days the maximum demand on breathing machines in hospitals occurs on April 18th, 2020 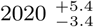 days. The first pandemic wave ends in Germany end of May 2020. The predictions are based on the assumption of a Gaussian time evolution well justified by the central limit theorem of statistics. The width and the maximum time and thus the duration of this Gaussian distribution are determined from a statistical *χ*^2^-fit to the observed doubling times before March 28, 2020.

## I. INTRODUCTION

In these days there is a very high interest in the societal, economical and political world to understand the time evolution of the first wave of infections of the population by the current Sars-Cov-2 (corona) virus. The most important issues are the total duration and the peak time of the infection evolution as well as the maximum number of daily infections. It would be most helpful for many people to have a reproducable, crude, but reliable estimate when this pandemic wave is over. It is the purpose of this manuscript to provide such an estimate based on a simplified Gaussian model for the time development of the pandemic outburst.

The best justification for the Gaussian or normal distribution for the virus time evolution is given by the central limit theorem of statistics^1^. The central limit theorem states that in situations, when many *n* ≫ 1 independent random variables are added, their properly normalized sum tends toward a normal or Gaussian distribution function of the form (1) even if the original variables themselves are not normally distributed. The spread of the virus infection of populations with high number of persons certainly is such a random process to which the central limit theorem ia applicable. Each person in a given population has a probability distribution (normalized to unity) as a function of time of being infected: it is a very noncontinuous distribution being 1 at the day of infection and 0 on all other days. If one adds up these discrete distributions of persons living in villages and districts of towns of typical size of about 1000 persons one obtains quasi-continous probability distributions for being infected which certainly will be different in hotspots of the disease and isolated rural areas. If we then add up a large number of these village probability distributions for all of Germany we obtain the daily infection rate distribution which according to the central limit is close to a Gaussian distribution.

The analysis of Gaussian distribution functions plays a central role in many problems of statistical physics and plasma physics. E.g. in plasma kinetic theory they are referred to as drift-Maxwellian^2^ or counterstreaming bi-Maxwellian^3,4^ velocity distribution functions. Both authors of this manuscript are not virologists but theoret*ical* physicists in plasma physics and astrophysics (RS) and solid state physics (FS) with past experience in analyzing normal or Gaussian distribution functions.

Apart from consulting several reviews^5–7^, as non-virologists we are not familiar with the recent relevant virology literature. Nevertheless, it is our hope that in these hard times an estimate by unbiased non-experts might be welcomed by specialists as well as the broad population, especially if some positive information and outlook is provided. We base our parameter estimates on publicly available information, especially by the excellent podcast by Prof. C. Drosten^8^ and the recent sophisticated modelling study for Germany^7^.

## II. GAUSSIAN MODEL

Numerical simulations and the empirical data of earlier epidemies^8^ indicate that the time evolution of epidemic waves is characterized by an early exponential rise until a pronounced maximum is reached followed by a rapid decrease. As argued above we adopt a simple Gaussian model for the time evolution of infections and explore its consequences. If *I*(*t*) denotes the number of infections per day, we assume that its time evolution is given by the Gaussian function

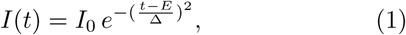

shown in Fig. 1, where *I*_0_ denotes the maximum value at time *E* and Δ denotes the width of the Gaussian.

**FIG. 1.**
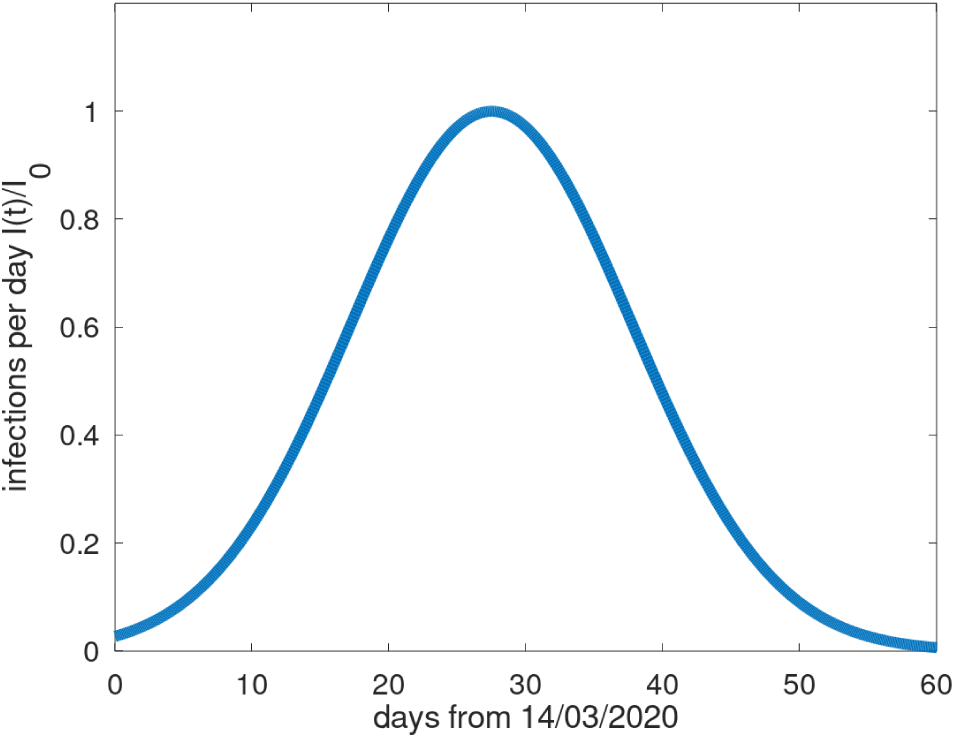
Prediction of the time evolution *I*(*t*)*/I*_0_ for Germany for the best fit parameters *E* = 27.5 days and Δ = 14.4 days. The time *t* = 0 corresponds with the date March 14, 2020, the day monitoring of the doubling times and drastic political actions against the virus started.

By monitoring the new daily infections one easily derives the relative change in the infections per day

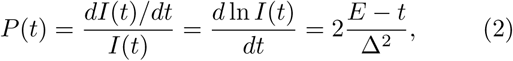

where we used the distribution (1).

### A. Doubling time

The monitored data are often given in terms of the doubling time *d* of the corresponding exponential function at any time

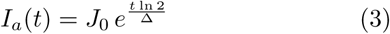

Using the distribution (3) in equation (2) provides for the relative change in daily infection rate

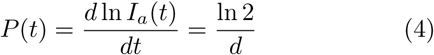

Equating the two results (2) ans (4) leads to the time-dependent Gaussian doubling time

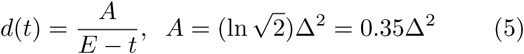

Figure 2 shows the monitored doubling times for Germany^9^ starting on March 15, 2020 until March 28,2020. We assume that every value has an error of 15 percent. It starts at *d*(*t* = 0) = 2.6 days and increases to *D*(*t* = 12) = 4.8 ± 0.24 on March 28, 2020.

**FIG. 2.**
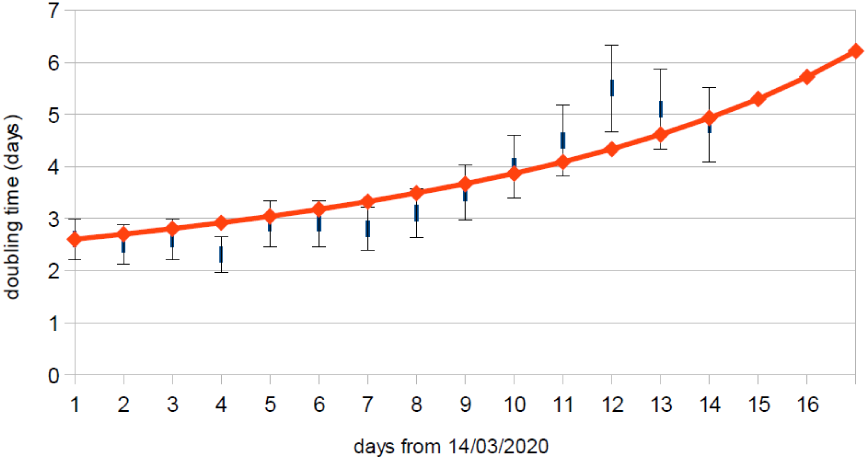
Monitored doubling times in Germany from March 15 until March 28, 2020 in comparison with the best fit

The Gaussian doubling time modeling then provides *d*(*t* = 0) = *A/E* = 2.6 days, corresponding to

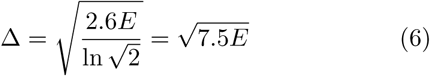

ln Moreover, Eq. (5) reduces to

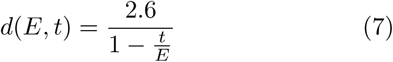

### B. Statistical fit

We determine the value of the only free parameter *E* in equation (7) by performing a *χ*^2^-fit to the data shown in Fig. 2. If *m*(*t*_*i*_) denote the observed doubling times at days *t*_*i*_, *δm*(*t*_*i*_) = 0.15*m*(*t*_*i*_) its error and *d*(*E, t*_*i*_) the theoretical doubling time for given values of *E*, we calculate

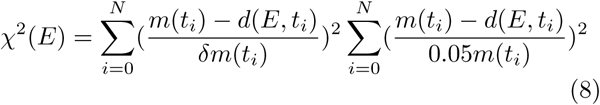

Currently, on March 28, 2020 we have *N* = 14 measurements. With one free parameter the number of degrees of freedom is *N* − 1 = 13.

The best fit with the minimum value of 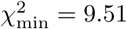 is provided for *E* = 27.5 days. The 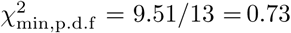 per degree of freedom is less than unity indicating that our model fits the data very well. For 90 percent confidence all values of 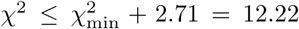 are acceptable^10^ yielding with 90 percent confidence that

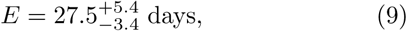

correponding to April 11, 2020 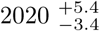 days. Consequently, the best fit Gaussian doubling time for Germany is given by

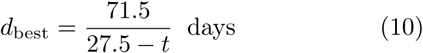

As an aside we note that the variation (10) becomes infinitely large as *t* → 27.5. Moreover, for times *t > E* the doubling times becomes a decay half-life approaching 0 for very large times *t* ≫ *E*.

Moreover, after inserting the values (9) equation (6) yields with 90 percent confidence

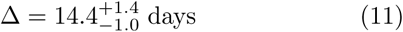

In Fig. 3 we show the prediction of the doubling times in Germany until day 25 corresponding to April 8th, 2020.

**FIG. 3.**
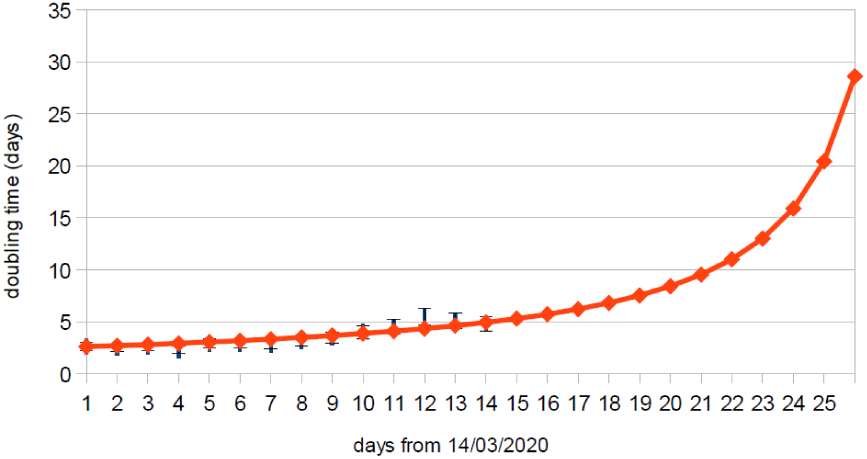
The same as in Fig. 2 but now the predicted doubling times until day 25 corresponding to April 8, 2020.

## III. PREDICTIONS FOR GERMANY

### A. Total number of infections

It is known that during the whole duration of the first wave of the virus evolution 70 percent of the total population are infected^8^, if nothing is done to reduce the number of infections. Scaling the total population in units of 10^5^*N*_5_, we estimate that 0.7*q*10^5^*N*_5_ are infected during the whole duration of the first virus wave, where the quarantaning factor *q* accounts for the currently taken political actions such as quarantining of elder and infected people, social distancing actions in the society as well as the closure of schools and daycare facilities.

Integrating the Gaussian (1) over all times we then obtain

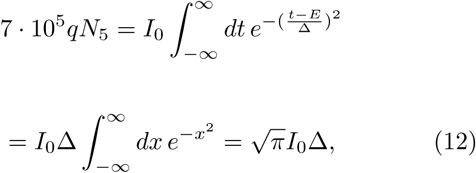

where we used the integral 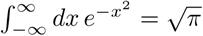. Equation (12) yields for the maximum value

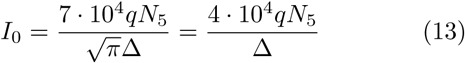

With the 90 percent confidence value for Δ from equation (11) we obtain with the same confidence level for the maximum value (13)

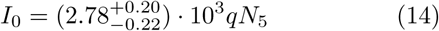

### B. Manageable infections

We assume as typical medical parameters those suggested by the recent modeling by the Robert-Koch-Institute^7^ (see their Fig. 1): only about 20 percent of the infected people are seriously infected, 5 percent have to be hospitalized and 1*α* percent need access to breathing apparati for typically 1*w* week, corresponding to 7*w* days. We refer to the latter as NSSPs standing for new seriously sick persons per day. As these numbers are uncertain we keep their scalings with the typically adopted numbers.

In Germany there are at most 56000 breathing apparati in total available, corresponding for a population of 80 million people to 70*b*_70_ breathing apparati available per 10^5^*N*_5_ people. As a consequence, every day the hospitals can handle *I*_*n*_ NSSPs with

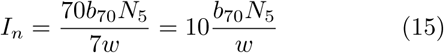

The infections are handable by hospitals for all times if at the maximum of the virus evolution 0.01*αI*_0_, denoting the maximum number of seriously sick persons per day needing access to breathing apparati, is less or equal to *I*_*n*_, i.e.

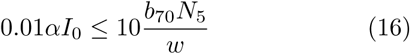

Inserting the value (14) for the maximum value *I*_0_, the quantity *N*_5_ cancels out and we obtain with 90 percent confidence the condition

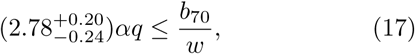

which is equivalent to

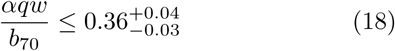

In order to handle all serious infections in German hospitals the condition (18) has to be fulfilled. It seems that German hospitals can only ensure the best treatment of all NSSPs at the maximum of first wave if either (1) the number of available breathing apparati can be increased by a factor of 3, corresponding to *b* = 210 per day. or (2) the quarantaining factor *q* can be reduced to 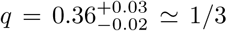 The first option is unrealistic on short time scales. To achieve the second option of the reducing the quarantaining factor *q* to about 0.3 by the currently taken social distancing and quarantaining activities seems to be realistically achievable in Germany. We therefore will adopt this optimistic value *q* = 0.3*q*_0.3_ in our further predictions. However we note that with such a small quarantaining factor only 21000*N*_5_ will be infected during the whole first wave of the virus, so that additionally waves are likely to occur in the future.

It is important to notice that the outbreak of serious sickness syndroms of NSSPs is delayed to the infection time by about *τ* = 7 days^8^. This delay time has to be added to the above derived maximum time scale *E*, so that 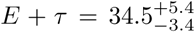 days, corresponding to April 18, 2020 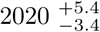 is the predicted day in Germany when the maximum number of NSSPs has to be treated.

### C. Duration of the first wave

The number of infections are signifantly reduced by a factor 10^3^ compared to the maximum *I*_0_ at the time

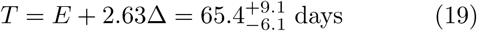

with 90 percent confidence. Consequently, the first pandemic wave will be over in Germany not before 65 days or about 2 months with the indicated uncertainty. This corresponds to 38 ± 4 days after the time of maximum.

According to our predictions, the first pandemic wave in Germany will reach its maximum by April 18, 2020 when about about (8.3 ± 0.78)*q*_0.3_*N*_5_ NSSPs have to be treated in the hospitals. The wave has a broad distribution from April 6 to April 30, 2020 with more more than (4.2 0 ± 39)*q*_0.3_*N*_5_ but less than (8.3 ± 0.78)*q*_0.3_*N*_5_ NSSPs per day. The number of NSSPs needed to be treated at hospitals will sharply drop to less than (0.0083 ± 0.00078)*q*_0.30_*N*_5_ NSSPs by May 26th, 2020.

As Germany has a population of about 80 million persons we have *N*_5_ = 800. Therefore in absolute numbers German hospitals will have to cope with (6640 ± 624)*q*_0.3_ NSSPs at the maximum of the outburst on April 18th, 2020, more than (3360 ± 312)*q*_0.3_ but less than 6640 ± 624 NSSPs per day between April 6 and April 30, 2020, before the total number per day drops below (6.64 ± 0.63)*q*_0.3_ NSSPs after the end of May 2020. All errors have 90 percent confidence.

Our analysis can be applied to other countries too if reliable information on the early doubling times are available. We plan to test our modeling with the data from the past first corona wave in China.

### D. Final important remark

We end with a sentence of caution: although the central limit theorem provides us with a very good justification of the adopted Gaussian time distribution function it is not guaranteed that the actual virus time evolution follows this behavior. We will only know for sure after the first pandemic wave is over. So it is possible that our estimates and we are wrong. We take this risk because we are convinced that many persons will welcome our optimistic estimate that the first wave is over by end of May 2020. There is light at the end of a long tunnel. Our estimate might also help decision makers when to lift the current societal and economical lockdown.

## Data Availability

All used data are publically available

## IV. DEUTSCHE ZUSAMMENFASSUNG: CORONA IN DEUTSCHLAND VORBEI ENDE MAI?

Mit heutigen Datum 30. März 2020 wird vorhergesagt, dass mit 90-prozentiger Konfidenz die erste Welle der Corona-Pandemie in Deutschland die maximale Zahl von neuinfizierten Personen am 12. April 2020 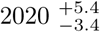 Tage erreicht. Mit einer Verzögerungszeit von etwa 7 Tagen wird dann am 19. April 2020 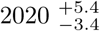 Tage der höchste Ansturm auf Beatmungsgeräte in den Krankenhäusern erfolgen. Die erste Welle wird gegen Ende Mai 2020 in Deutschland enden mit einem tausendstel kleineren Neuinfektionsraten. Diese Vorhersagen basieren auf der Annahme einer Gauss-förmigen Zeitverteilung der Infizierungsrate, die gut durch den zentralen Grenzwertsatz der Statistik begründet ist. Die Breite und die Zeit des Maximums der Gauss-Verteilung und damit deren Gesamtdauer werden durch einen statistischen *χ*^2^-Fit an die in Deutschland beobachteten Verdopplungszeiten vor dem 28.März 2020 bestimmt. Die Behandlung aller schwer erkrankten Patienten mit Beatmungsgeräten ist gewährleistet, wenn es durch die andauernden Quarantäne- und soziale Distanzierungs-Massnahmen gelingt, die Anzahl der infizierten Personen in der Bevölkerung durch die erste Welle unter 30 Prozent zu halten.

